# Exploring the ATN classification system using Brain Morphology

**DOI:** 10.1101/2022.06.01.22275839

**Authors:** Nils Heinzinger, Anne Maass, David Berron, Renat Yakupov, Oliver Peters, Jochen Fiebach, Kersten Villringer, Lukas Preis, Josef Priller, Eike Jacob Spruth, Slawek Altenstein, Anja Schneider, Klaus Fliessbach, Jens Wiltfang, Claudia Bartels, Frank Jessen, Franziska Maier, Wenzel Glanz, Katharina Buerger, Daniel Janowitz, Martin Dichgans, Robert Perneczky, Boris-Stephan Rauchmann, Stefan Teipel, Ingo Killimann, Doreen Göerß, Christoph Laske, Matthias H. Munk, Annika Spottke, Nina Roy, Michael T. Heneka, Frederic Brosseron, Laura Dobisch, Michael Ewers, Peter Dechent, John Dylan Haynes, Klaus Scheffler, Steffen Wolfsgruber, Luca Kleineidam, Matthias Schmid, Moritz Berger, Emrah Düzel, Gabriel Ziegler

**Author notes:** Shared last-authorship (Emrah Düzel, Gabriel Ziegler). Correspondence to: Nils Heinzinger; Institute for Cognitive Neurology and Dementia Research; University Hospital Magdeburg, University of Magdeburg, Leipziger Str. 44, 39120 Magdeburg, Germany.

## Abstract

**Background:** The NIA-AA proposed Amyloid-Tau-Neurodegeneration (ATN) as a classification system for AD biomarkers. The Amyloid Cascade Hypothesis (ACH) implies a sequence across ATN groups that patients might undergo during transition from healthy towards AD: A-T-N-→A+T-N-→A+T+N-→A+T+N+. Here we assess the evidence for monotonic brain volume decline for this particular (Amyloid-conversion first, Tau-conversion second, N-conversion last) and alternative progressions using Voxel-based Morphometry (VBM) in a large cross-sectional MRI cohort.

**Methods:** We used baseline data of the DELCODE cohort of 437 subjects (127 Controls, 168 SCD, 87 MCI, 55 AD patients) which underwent lumbar puncture, MRI scanning and neuropsychological assessment. ATN classification was performed using CSF-Aβ42/Aβ40 (A+/-), CSF-phospho-Tau (T+/-), and adjusted hippocampal volume or CSF-total-Tau (N+/-). We compared voxel-wise model evidence for monotonic decline of gray matter volume across various sequences over ATN groups using the Bayesian Information Criterion (including also ROIs of Braak stages). First, face validity of the ACH transition sequence A-T-N-→A+T-N-→A+T+N-→A+T+N+ was compared against biologically less plausible (permuted) sequences among AD-continuum ATN groups. Second, we evaluated evidence for 6 monotonic brain volume progressions from A-T-N-towards A+T+N+ including also non-AD-continuum ATN groups.

**Results:** The ACH-based progression A-T-N-→A+T-N-→A+T+N-→A+T+N+ was consistent with cognitive decline and clinical diagnosis. Using hippocampal volume for operationalization of neurodegeneration (N), ACH was most evident in 9% of gray matter predominantly in the medial temporal lobe. Many cortical regions suggested alternative non-monotonic volume progressions over ACH progression groups, which is compatible with an early amyloid-related tissue expansion or sampling effects e.g. due to brain-reserve. Volume decline in 65% of gray matter was consistent with a progression where A status converts before T or N status (i.e. ACH/ANT) when compared to alternative sequences (TAN/TNA/NAT/NTA). Brain regions earlier affected by Tau tangle deposition (Braak stage I-IV, MTL, limbic system) present stronger evidence for volume decline than late Braak-stage ROIs (V/VI, cortical regions). Similar findings were observed when using CSF-total-Tau for N instead.

**Conclusion:** Using the ATN classification system, early Amyloid status conversion (before Tau and Neurodegeneration) is associated with brain volume loss observed during AD progression. The ATN system and the ACH are compatible with monotonic progression of MTL atrophy.

DRKS00007966, 04/05/2015, retrospectively registered

## Introduction

Alzheimer’s disease (AD) is a slowly evolving neurodegenerative condition where initial brain changes can be found up to decades before the clinical onset and ultimately result in progredient cognitive decline and brain atrophy often studied with Magnetic Resonance Imaging (MRI)^1-3^.

AD is characterized by the accumulation of protein deposits, i.e. β-Amyloid plaques and neurofibrillary tangles (NFT) consisting of hyperphosphorylated Tau which can be assessed using cerebrospinal fluid (CSF) biomarkers^4,5^. A reliable marker reflecting Amyloid deposition is the Aβ42/Aβ40 ratio which decreases with increasing deposition^6^. Accumulation of Tau tangles is mirrored by increasing CSF hyperphosphorylated Tau, while CSF total Tau has been more generally associated with neuronal loss, not necessarily AD specific. Those biomarkers have shown potential for predicting the clinical diagnostic conversions^7-9^ and worsening of memory performance during disease progression^10^.

One key concept about the disease progress and pathological timeline has been introduced as the Amyloid Cascade Hypothesis (ACH)^11-13,1^. Due to different predispositions, including age^14^, genes^15^ or vascular risk factors^16^, ß-Amyloid is increasingly formed from precursor proteins which leads to its aggregation in the brain. Then, ß-Amyloid can induce hyperphosphorylation and malformation/misfolding of intracellular Tau proteins, which aggregate in forms of NFTs^13^. Increased cellular stress results in neuronal loss which typically manifests behaviourally in progressive cognitive decline. Neuronal death in AD manifests in a typical MRI atrophy pattern with strongest morphometrical changes situated in medial temporal lobe (MTL) and other limbic regions, while the primary motor and sensory cortex are often spared^17,18^. Although the ACH was postulated about 30 years ago, the hypothesis is still under refinement and critical review^13,19,20^. Moreover, the stereotypical progression pattern of Tau/NFT spread from the transentorhinal region via the limbic system to the whole cortex during AD progression can be classified into six Braak stages, which have been first described in an autopsy study^5^, and later tested in positron emission tomography studies^21,22^ or VBM atrophy studies^23^.

Recently, a new descriptive ATN classification for AD which emphasises pathological and physiological rather than traditional clinical measures such as neuropsychological test scores was proposed^24,25^. In the ATN system, for the three binary categories Amyloid burden, Tau burden and Neurodegeneration, subjects are rated as normal (physiological, “-”) or abnormal (pathological, “+”). The resulting 8 (=2^3^) groups with different biomarker combinations range from A-T-N-(suggesting no pathology) to A+T+N+ (with pathology in all categories). It has been suggested that all ATN biomarker combinations with A+ reflect a pathological change related to the AD continuum. Several recent studies explored the prognostic possibilities for clinical progression and cognitive decline using ATN^26-30^. However, while the ATN classification does not directly imply a progression cascade or a set of subsequently following stages per se, it may be used for this particular purpose. For example, the sequence of a disease transition across pathology groups (1) A-T-N-(2) A+T-N-(3) A+T+N-(4) A+T+N+ is more compatible with the amyloid cascade hypothesis than other progression sequences based on ATN classification groups^25^. If individual participants follow this particular disease progression profile, this would imply a monotonic volume loss across groups (1)→(2)→(3)→(4) in brain areas associated with AD. While above progression sequence is partially supported in selected studies^31,32^, those findings are limited to recordings of non-imaging between-group biomarker differences. Although there is evidence for deviating sequences of progression^32^, studies focusing on local voxel-based anatomical analysis in relation to ATN groups are still missing (see eg.^33^).

Here we study whether above progression implied by the ACH is reflected in specific patterns of local GM volume decline using cross-sectional data from a large neuroimaging cohort (DELCODE; DZNE Longitudinal Cognitive Impairment and Dementia Study) which is well characterized by CSF biomarkers. The DELCODE cohort is specifically enriched in subjects that are at risk for developing AD such as Subjective Cognitive Decline (SCD), but also Mild Cognitive Impairment (MCI) and thus more likely comprises individuals in early preclinical stages of AD (A+).

GM volume is a sensitive marker for local brain changes or pathological processes. As this marker is continuous, smallest substance differences for all brain regions can be measured and intermediate changes are detectable even when they would not cause an ATN status conversion. We hypothesize that GM in the hippocampal-network decreases following the ACH sequence and (1) test face validity of an ACH-based sequence using Voxel-based Morphometry without *a-priori* regional assumptions; and (2) compare the evidence for volume loss reflecting the ACH sequence in comparison to other biologically possible progressions outside the AD continuum. Finally, the concordance between ACH progress and Braak staging is evaluated. We expect earlier Braak stages to be stronger affected by atrophy during the ACH sequence. It might occur that the volume alteration is regionally modulated by e.g. reserve mechanisms. Since operationalization might be crucial, we also evaluate the impact of alternative choices for dichotomization of the N category using both t-Tau or hippocampal volume.

## Methods

### Study Design and Participants

This study uses the baseline data of the DELCODE cohort, an observational multicentre study with 10 sites from the German Centre of Neurodegenerative Diseases (DZNE). Its focus is the multimodal assessment of preclinical stages of dementia of Alzheimer’s type (DAT) including SCD, MCI, DAT, and DAT relatives^34^. While SCD, MCI and DAT participants were recruited from memory clinics, relatives of DAT patients and healthy controls were recruited by advertisement and initially screened per phone for self-experienced cognitive decline and memory worries. Further SCD inclusion criteria were a normal cognitive performance (specified as within 1.5 SD compared to an age, sex and education years adjusted control group) in all subtests of the CERAD-plus battery and a MMSE score between 26 and 30 and a CDR score <= 0.5.

Participants with MCI were below 1.5 SD in the CERAD-plus battery, but did not fulfil dementia criteria of NINDCS/ADRDA^35^. Subjects diagnosed as DAT were fulfilling NINDCS/ADRDA criteria, have a CERAD-plus score of below 1.5 SD, and were within an extended MMSE score range of 18-26 and have a CDR rating of >= 1. DAT relatives have a first-grade sibling with diagnosed DAT and do not fulfil MCI or DAT criteria.

Noncomplaining healthy controls (NC) neither suffered from subjective or objective cognitive impairment. All participants were native German speakers, older than 60 years, and gave written informed consent and had a study partner available for consultation. Other neurological or psychiatric disorders than DAT were excluded. More information on study design and inclusion/exclusion criteria can be found elsewhere^34^. DELCODE is retrospectively registered at the German Clinical Trials Register (DRKS00007966), (04/05/2015) and was approved by ethical committees and local review boards. Of a total of 1079 participants at baseline timepoint, we finally included 437 subjects with available quality checked MRI imaging and CSF biomarkers (see below). Based on a clinical classification approach, this includes 127 NC (including DAT relatives), 168 SCD, 87 MCI and 55 DAT patients. A summary of demographic information of the analysed sample is provided in results table 1.

**Table 1:**
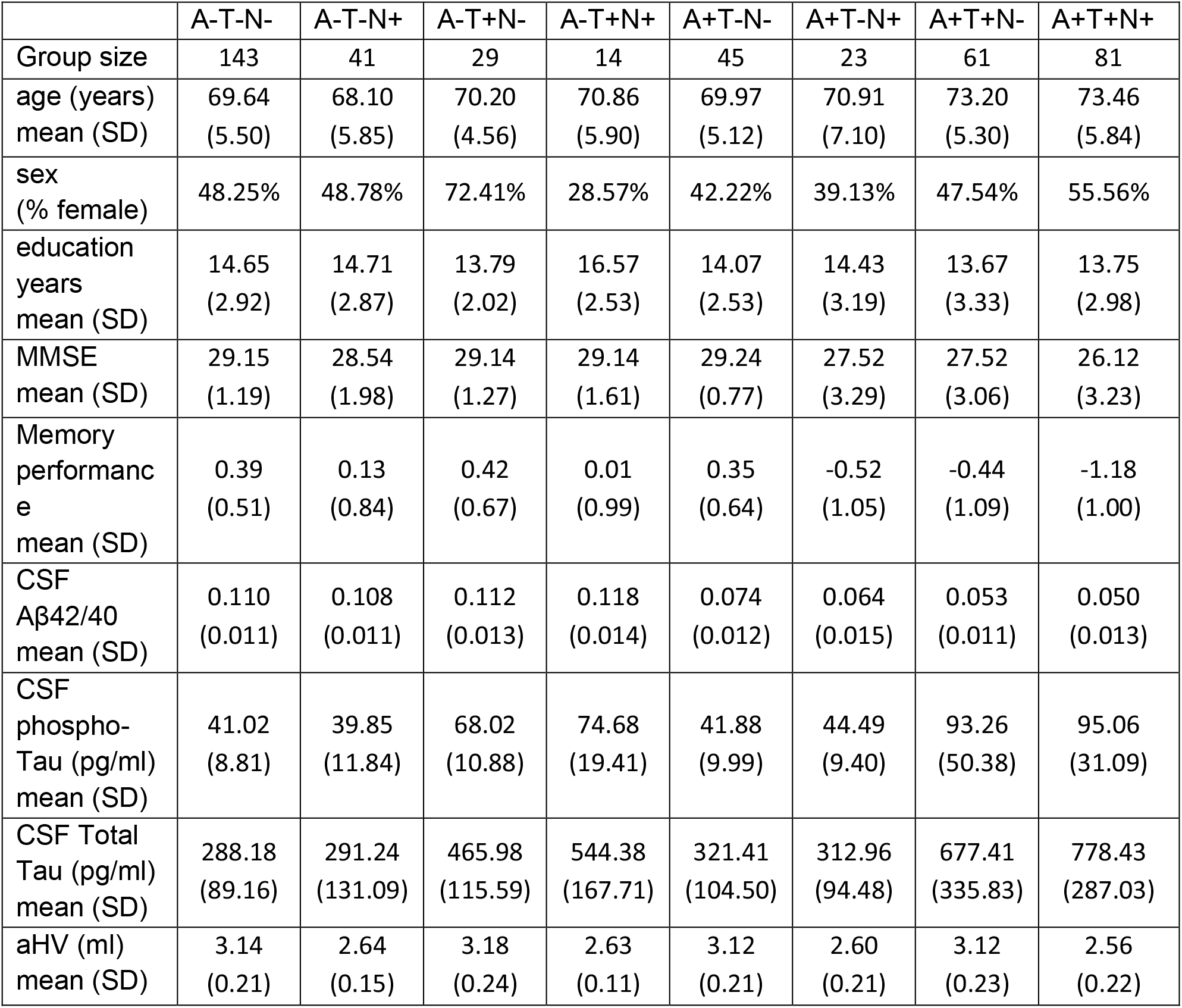
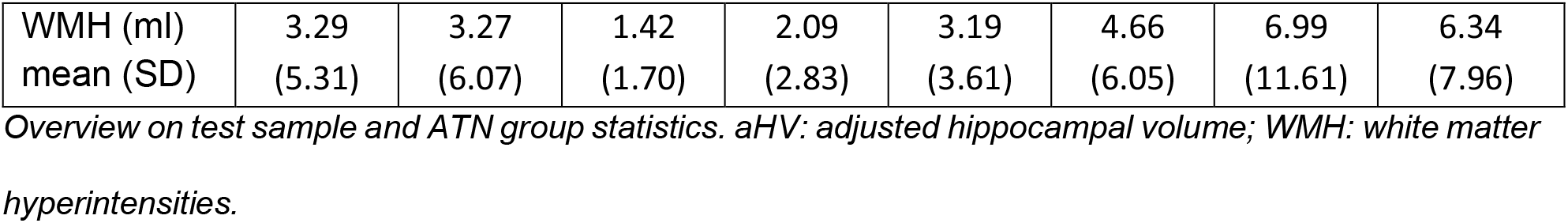
Sample and ATN group overview.

### Neuropsychological testing

In DELCODE, subjects underwent a large battery of neuropsychological tests. Due to our focus on global cognition and memory aspects in healthy and (pre-) clinical DAT patients, we use the Mini Mental State Examination (MMSE, ^36^) and a reliable memory composite factor score (further denoted as memory performance). This score was created by Confirmatory Factor Analysis and enables detecting subtle cognitive deviations in SCD when compared to NC subjects^37^.

### Biomarker and MRI data acquisition

Lumbar puncture was carried out by trained study assistants in 49% of DELCODE participants. CSF samples were centrifuged, aliquoted and stored at -80°C for retests. Biomarkers known to be related to AD pathology (CSF Aβ42, Total Tau, hyperphosphorylated Tau) were determined by commercially available kits (V-PLEX Aβ Peptide Panel 1 (6E10) Kit (K15200E), V-PLEX Human Total Tau Kit (K151LAE) (both Mesoscale Diagnostics LLC, Rockville, USA), Innotest Phospho-Tau(181P) (81581; Fujirebio Germany GmbH, Hannover, Germany)).

MRI scans were acquired in 9 out of 10 involved DZNE sites (3T Siemens scanners: 3 TIM Trio systems, 4 Verio systems, 1 Skyra and 1 Prisma system). Our main analyses were based on whole brain T1-weighted MPRAGE (3D GRAPPA PAT 2, 1 mm^3^ isotropic, 256 × 256 px, 192 slices, sagittal, ∼5 min, TR 2500 ms, TE 4.33 ms, TI 110 ms, FA 7°). Further ROI and covariate processing was based on additionally available FLAIR and T2-weighted protocols (for details see^34^). Additional details on standard operation procedures, quality assurance and assessment (QA), performed by the DZNE imaging network (iNET, Magdeburg), can be found elsewhere^34^.

### Image processing and computational brain morphometry

The MPRAGE images were processed using SPM (SPM12 v7771, Statistical Parametric Mapping software; Wellcome Trust Centre for Human Neuroimaging, London, UK, ^38^) and CAT-Toolbox (CAT12.6 r1450, Structural Brain Mapping group, Jena University Hospital, Jena, Germany, ^39^) under MATLAB (r2019b, The MathWorks, Inc., Natick, Massachusetts, USA). As first step, a correction for field inhomogeneities was applied. Then the images were segmented into GM, WM and CSF maps using CAT which includes a partial volume estimation correction on AMAP approach^40^. The received tissue maps with a 1 mm isometric voxel size are warped to a study-specific template in MNI space using Geodesic Shooting approach^41^. The GM tissue maps were modulated by the jacobian determinant to enable voxel-based comparisons of local gray matter volume across subjects. A Gaussian blurring kernel was applied with 6 mm full width half maximum (FWHM). The resulting tissue maps were quality tested using CAT’s sample homogeneity check and 15 subjects were excluded due to preprocessing artifacts.

For complementary ROI analysis we used Freesurfer’s (v6.0, ^42^) volume reconstruction (cortical stream^43^, subcortical stream^44^) to extract region of interest volumes. This was carried out by the default pipeline initiated by a ‘recon-all -all’ command which contains all preprocessing steps needed, including for example intensity normalization, surface registration to Talairach space, skull stripping, subcortical segmentation and calculation of affiliated region statistics, WM segmentation, tessellation and inflation of pial parcellated WM surfaces and cortical parcellation with calculation of cortical region statistics. Four ROIs (amygdala, hippocampus, entorhinal cortex, precuneus), well known to be affected early by AD pathology, were assessed^45-47,17,48^. Furthermore, anatomical masks representing Braak stages were created following^49^ and warped to MNI space. Thus, the following cortical regions were included as aggregated volumes: stage I/II: entorhinal cortex and hippocampus, stage III/IV: limbic, insular and temporal regions, V/VI: remaining cortical regions including primary sensory/motor areas or precuneus.

In order to enable a reliable operationalization of the N category of ATN system we used the specifically developed hippocampal segmentation in Freesurfer that is based on a high-resolution T2-weighted scan of the medial temporal lobe^50^. Note, that the obtained hippocampal volumes were only used for the ATN classification of each participant, while all presented voxel-based and ROI volumes were based on conventional CAT and Freesurfer segmentations (as dependent variable). A strong co-occurrence of AD and white matter hyperintensities (WMH) as sign of vascular damage has been reported^16,51-54^. To account for WMH during our analyses, the total lesion volume was extracted using Lesion segmentation toolbox (v3.0.0, LPA and an 0.5 binary threshold, ^55,56^).

### ATN classification and group comparison

Each participant was classified as normal (-) or abnormal (+) in the Amyloid (A) and Tau (T) category depending on their biomarker levels of Aβ42over40 and phospho-Tau181 respectively. Cut-offs were estimated by a ROC analysis and Youden’s index (A = 0.09, T = 57 pg/ml; ^34^). In this study we explored effects of two different choices for the neurodegeneration (N) category. We focused on (1) adjusted hippocampal volume (denoted as aHV; cut-off = 2821.1 μl) and (2) CSF Total Tau (cut-off = 470 pg/ml). aHV was derived from the Freesurfer segmentation (see above) and corrected for age, sex, education, total intracranial volume (TICV) and WMH using a linear regression model. Dichotomization of participants’ aHV into N- and N+ was performed using Gaussian mixture modelling similar to established cut-off estimation for CSF biomarkers used for the A and T category^57^.

To assess group differences in age and education, one-way ANOVAs with ATN status as between-subject variable were used. Group differences in cognition (MMSE and memory performance) were tested in ANCOVAs with ATN status as between group variable and age, sex and education as covariates. In all cases, post-hoc analysis was performed by two sample t-tests using a Bonferroni correction to account for multiple comparisons. Notably, analyses were restricted to the four groups of the ACH-based progression (1) A-T-N-(2) A+T-N-(3) A+T+N-(4) A+T+N+ since we focused on implications for common AD-related trajectories. The distribution of ATN status per clinical diagnosis was tested by 2-sided Fisher’s exact test for distribution differences between cognitively unimpaired (NC, SCD) and cognitively impaired (MCI, DAT) subjects. Significance level is set to p < .05 (*) or p < .001 (**) respectively.

### Testing the evidence for a monotonic decrease of brain volume over ATN progression groups

We aimed to test the evidence of local brain GM volume loss as a process of AD progression. As predicted by the ACH, the volume would decline over the following groups (1) A-T-N-(2) A+T-N-(3) A+T+N-; to (4) A+T+N+. Thus, we hypothesized later ATN stages to be associated with significantly reduced GM in AD-related areas. We estimated a voxel-wise general linear model describing the local GM volume *y* for the 4 given groups as

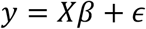

with design matrix *X*, coefficients *β* and residuals *ϵ*. The design matrix was chosen to define *β*_1_ as group mean of the first group, and for *g=2,3,4* coefficient *β*_*g*_ as group difference of group *g* and *g-1*. The model was fitted under linear constraints that *β*_*g*_ ≤ 0 for *g=2,3,4* and therefore implementing a monotonic decline of volume across groups *1* to *4* (using MATLAB R2019b’s function for constrained optimization lsqlin). For instance if a voxel has a true monotonic decline of volume *y* over groups *1,2,3,4* the model evidence is expected to be higher than for an alternative model with reversed group order e.g. *4,3,2,1*. We further compared different hypothesized and alternative sequences of volume decline progressions using the Bayesian Information Criterion^58^ (BIC) which compares the likelihood how well the data is described using a monotonic function while accounting for model complexity (Figure 1).

**Figure 1:**
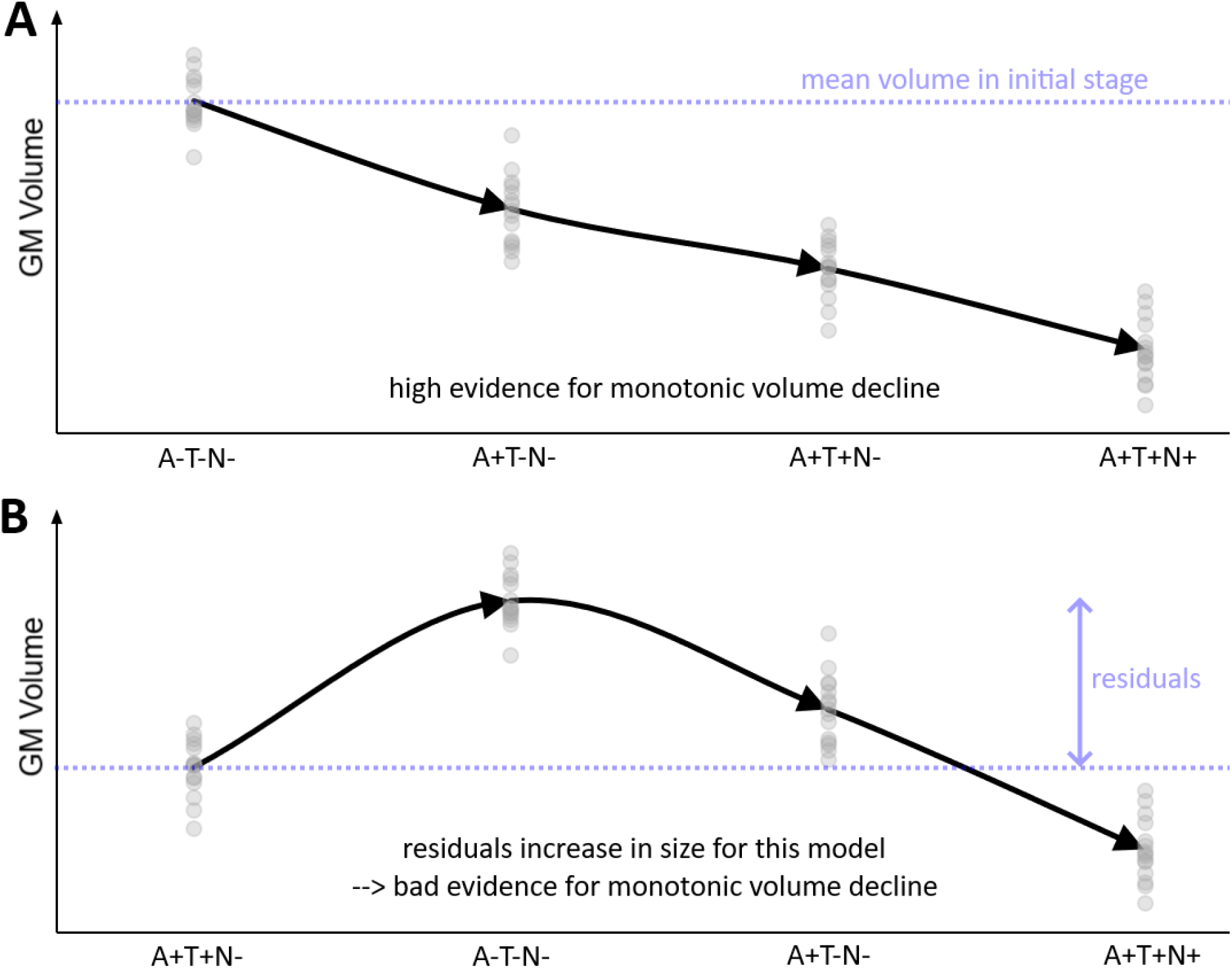
Monotonic and non-monotonic volume decline using ATN. A: An illustration of monotonic GM volume decline as hypothesised when following the ACH hypothesis using ATN groups. B: A permutated order of the upper case that clearly not shows a monotonic volume decline. A temporarily volume increase causes large residuals that cannot be explained by a monotonic model. Therefore, the pathway in A would be preferred over B (‘higher evidence for monotonic decline in A’).

First, face validity of the ACH hypothesis was tested by comparing the evidence of the above ACH-based sequence (1) → (2) → (3) → (4) against 23 (=4*3*2*1-1) alternative monotonic progressions generated by permutation which are *a-priori* less plausible if ACH is true. Note that this analysis was restricted to 4 primarily AD-related of all 8 possible ATN classification groups, where it is assumed that Amyloid conversion happens before status conversion of Tau and Neurodegeneration. In a second analysis, the evidence of the ACH-related volume trajectory was compared against 5 biologically plausible alternative sequences including also ATN groups which are considered outside the AD-continuum, in particular ANT (i.e. Amyloid-conversion first, N-conversion second, Tau-conversion last; therefore ‘ANT’), TAN, TNA, NAT, and NTA. These 6 sequences represent all conceivable possibilities to convert in three steps from A-T-N-(no pathology) to A+T+N+ (full pathology).

In this study, all tests were performed both on (A) whole brain voxel-based modulated GM volume images, and (B) a-priori hypothesized ROIs. Voxel-based tests were restricted to GM using an absolute threshold of .05. In addition, the percentage of voxels with the highest evidence for the ACH trajectory inside every ROI mask is provided.

All statistical analyses were performed in MATLAB. Voxel-wise test results are presented as maps with the highest evidence for one particular model and log p maps for inference on statistical parameters such as successive volume decline over groups (using FDR correction for multiple comparisons, p<0.05). Finally, the percentage of GM voxels with the highest evidence for a certain progression sequence is provided. All analyses were accounting for covariates age, sex, education, TICV and WHM. All main results are reported for N operationalized by aHV and selected results using CSF Total Tau can be found in the supplementary material.

## Results

### Sample demographics and ATN group comparisons

Key characteristics of the analysed sample are summarized in Table 1, selected comparisons (Bonferroni) can be found in Figure 2. As expected we found that age differed across ATN groups (F(7,429) = 6.65, p < .001). ATN groups showed also differences in years of education (F(7,429) = 2.63, p < .05). With respect to cognition, we found a significant effect of ATN group (F(7,426) = 13.46, p < .001), age (F(1,426) = 14.78, p < .001) and education (F(1,426) = 19.58, p < .001) on MMSE scoring, but no effect of sex (F(1,426) = 0.93, p = .33). Similar results were obtained for the memory performance, where ATN status (F(7,426) = 28.10, p < .001), age (F(1,426) = 44.54, p < .001), education (F(1,426) = 39.78, p < .001) and sex (F(1,426) = 3.87, p < .05) were significant.

**Figure 2:**
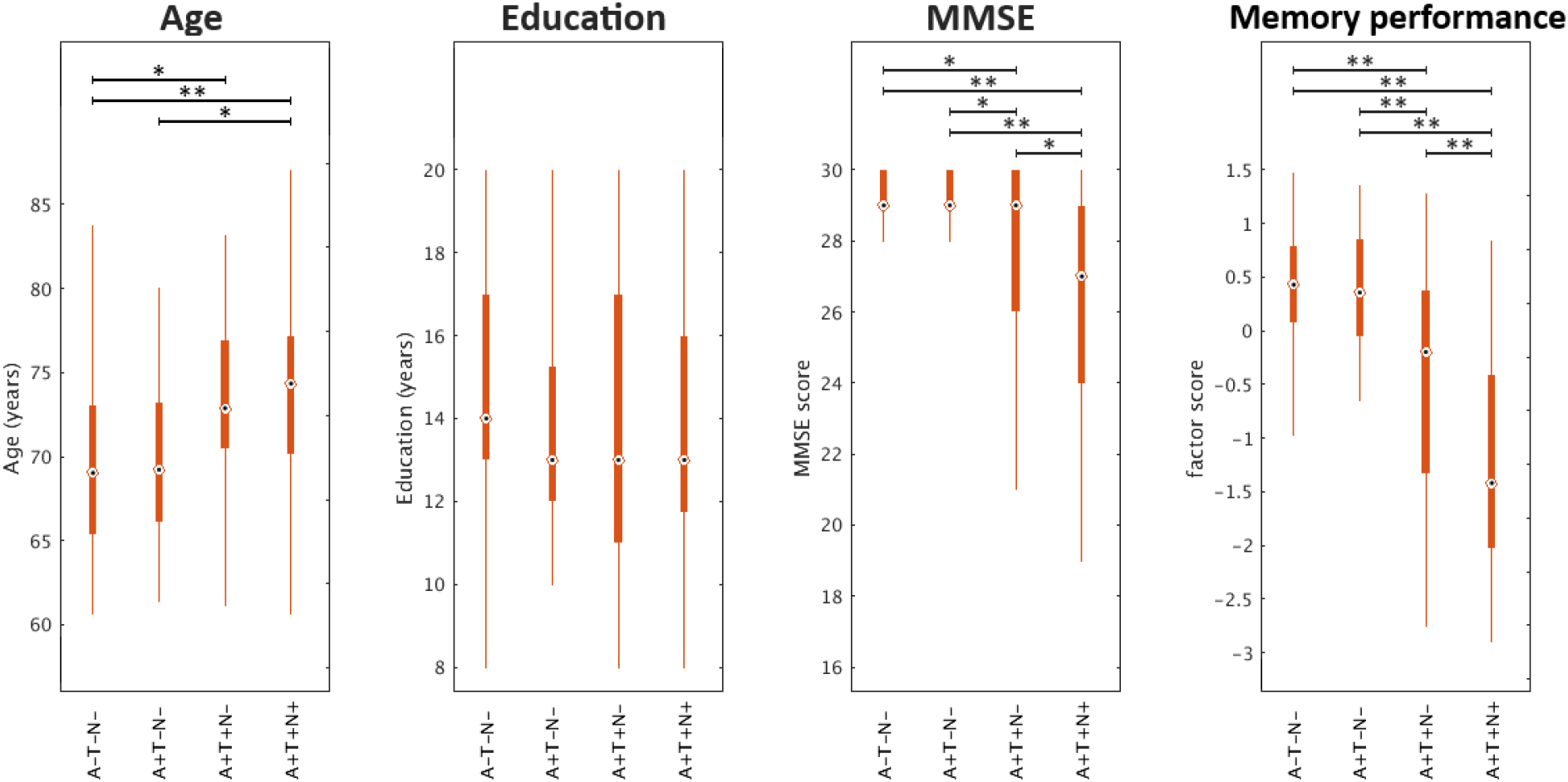
Comparison between selected ATN groups. Boxplots of age, sex, cognition for selected ATN groups. *: p < .05 after Bonferroni correction, **: p < .001 after Bonferroni correction.

As shown in Figure 2, the age increased while global cognition (MMSE) and memory performance decreased following a hypothesized disease progression using the ACH sequence (A-T-N-→ A+T-N-→ A+T+N-→ A+T+N+). No systematic pattern was found for years of education. These effects could be reproduced using CSF Total Tau for N (Supplemental File 1).

### Association of ATN status and clinical diagnosis

We observed an association of the ATN status and clinical diagnosis groups comparing cognitively unimpaired (NC, SCD) and cognitively impaired (MCI, DAT) participants for A-T-N-(p < .001), A-T+N-(p < .05), A+T-N-(p < .05), A+T+N-(p < .05) and A+T+N+ (p < .001). No non-random association was found for A-T-N+ (p = .49), A-T+N+ (p = .40) and A+T-N+ (p = .26). Compared to A-T-N-, the highest relative risk for DAT was found in A+T+N+ (30.90 times higher) and A+T-N+ (15.54 times). The lowest risk for DAT relative to A-T-N-was in A-T+N-(2.47 times higher), while no DAT cases were recorded in A-T+N+ or A+T-N-. Results suggested that more impaired clinical groups, especially DAT, were found more often among the Alzheimer’s continuum ATN groups (i.e. in A+), while cognitively unimpaired status was rather associated to no brain pathology (i.e. A-T-N-). For percentual distribution see Figure 3. A similar pattern was observed for N measured by CSF Total Tau (Supplemental File 2).

**Figure 3:**
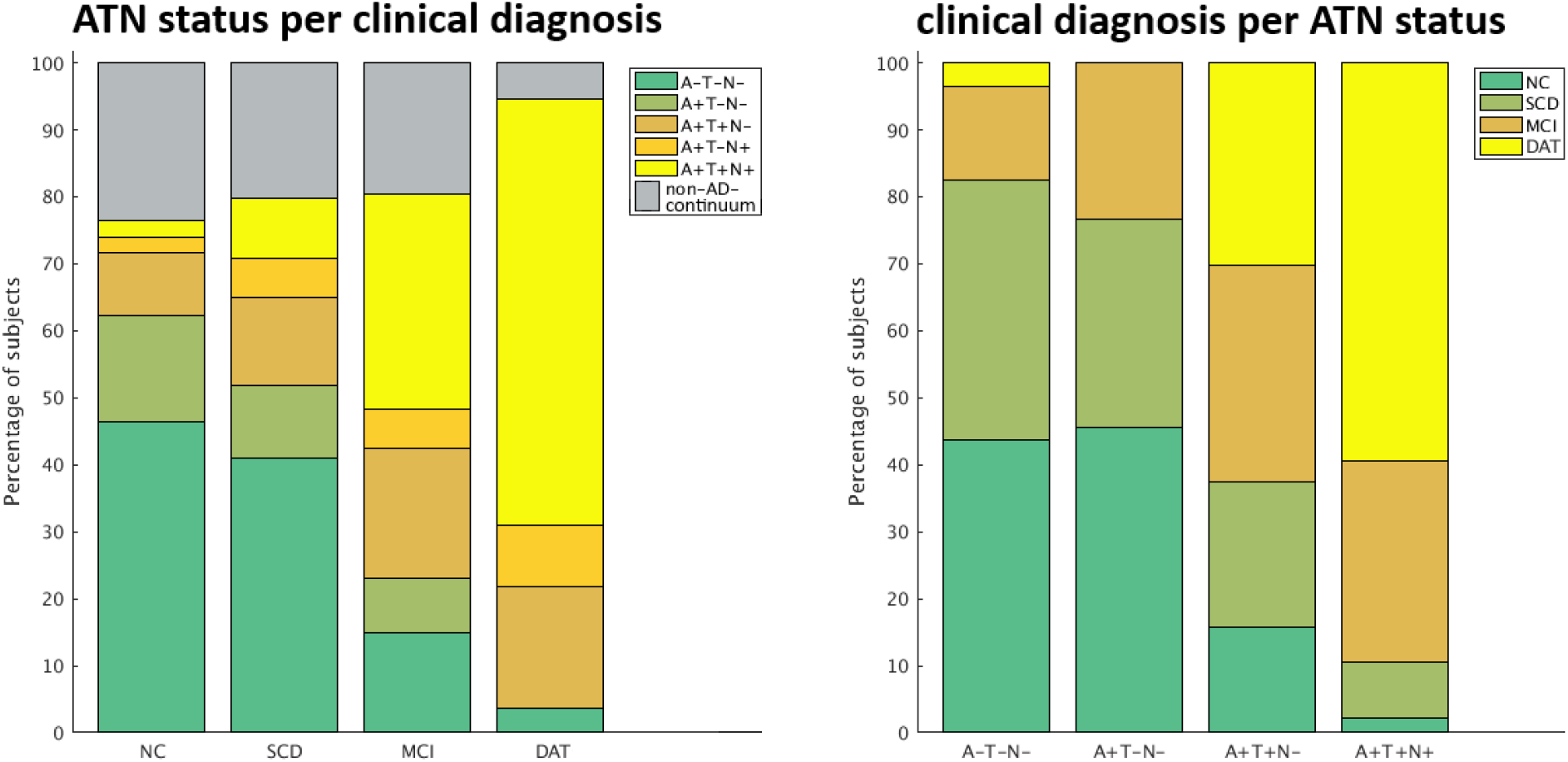
Distribution of ATN status and clinical diagnosis. Left: percentual distribution of selected ATN groups per clinical diagnosis; right: percentual distribution of clinical diagnosis per ATN groups.

### Assessing face validity of the ACH hypothesis using local brain volume

As a next goal, we identified brain regions where volume progression across ATN groups is compatible with the ACH hypothesis. More specifically, if the ACH is true it might be expected to observe a monotonic decline of volume over groups A-T-N-→ A+T-N-→ A+T+N-→ A+T+N+ in the hippocampal network^25^. The regions showing significant GM volume decline over this ACH sequence (of ATN biomarker conversions) are illustrated in Figure 4A (log p map, p < .05 FDR-corrected, N based on hippocampal volume, accounting for covariates age, sex, education, TICV and WMH). Strongest effects are found in the MTL region (peak: left post. hipp. x = -28, y = -22, z = -19, log p = 86.70). Further regions with significant GM volume loss following the ACH sequence are the orbital and basal forebrain, large parts of the temporal lobe, the insular cortex, the basal ganglia, the cingulate gyrus, the precuneus, (medial) premotor regions and the parietal and occipital lobes. When using CSF total Tau instead of hippocampal volume for operationalization of the ‘N’ category, we observed consistent but slightly less widespread shrinkage of local GM (peak: left ant. hipp. peak x = -26, y = -10, z = -17, log p = 21.41, Figure 4B).

**Figure 4:**
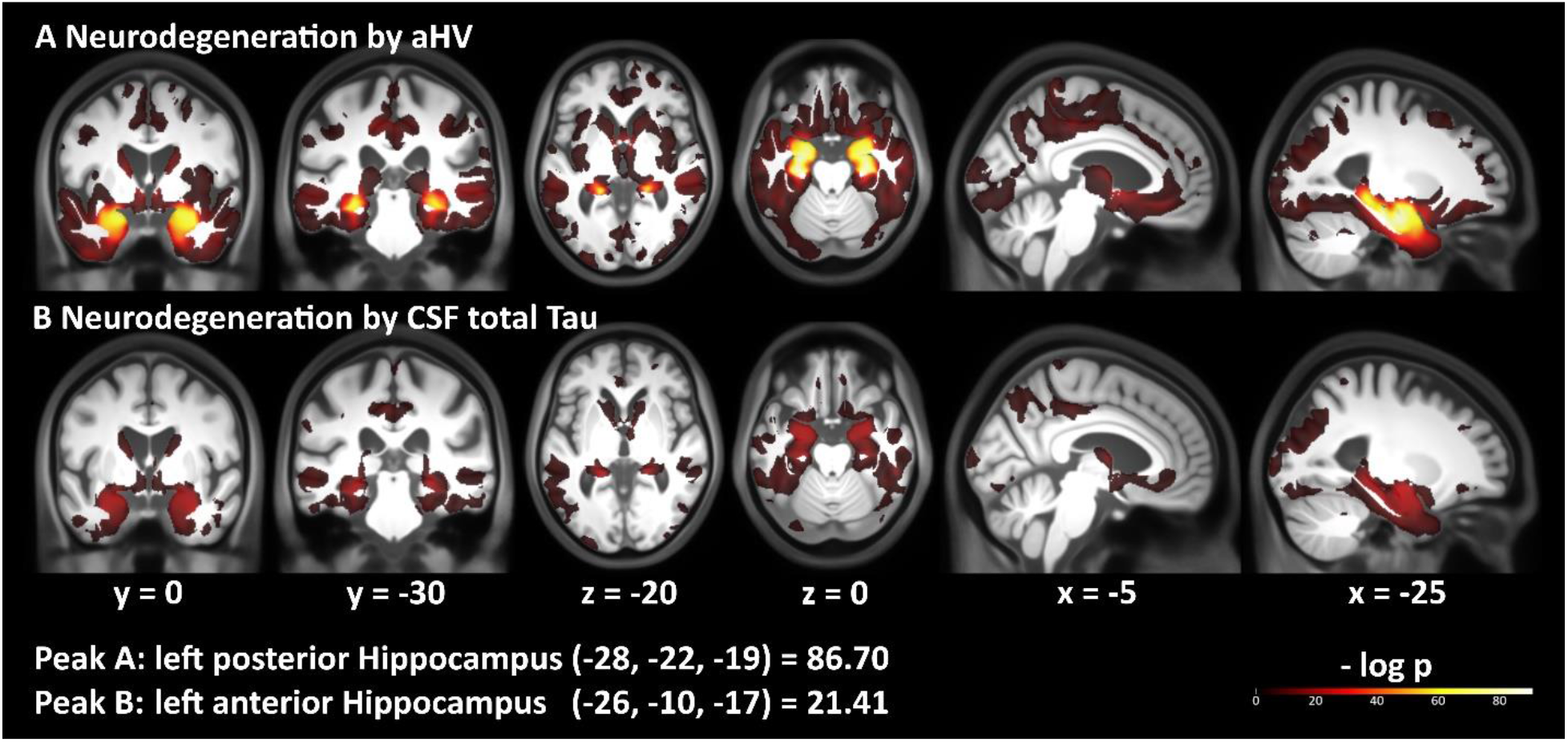
Volume decline following the ACH sequence. Regions showing significant GM volume loss along the ACH sequence. Unmasked log p map with p < .05, FDR-corrected. A: Neurodegeneration (N) defined by aHV; B: Neurodegeneration (N) defined by CSF total Tau.

It is important to note that testing for ‘any’ local volume decline over groups that align with the ACH-related progression might still reveal brain areas where alternative disease progressions are even more likely. Therefore, in an explorative analysis we compared voxel-wise evidence of the hypothesized ACH progression (or model) against 23 biologically less plausible (permuted) conversion sequences among the ATN classification groups associated with the AD continuum e.g. the above stated group progression but in reversed order. First, we applied a voxel-based test of monotonic GM volume decline using the Bayesian Information Criterion. Figure 5a illustrates the resulting regions with highest evidence for three selected progressions. Since only one sequence of diagnostic conversions can have the highest evidence in a given brain region (when compared to other progressions), these maps revealed non-overlapping areas of the brain. For 8.99% of all explored GM brain regions, ACH was indeed found to be the most evident progression sequence showing monotonic volume decline (Figure 5b). This especially involved the anterior MTL, hippocampus, parahippocampal gyrus and fusiform gyrus while the general pattern of regions most compatible with ACH is similar to the above presented findings.

**Figure 5:**
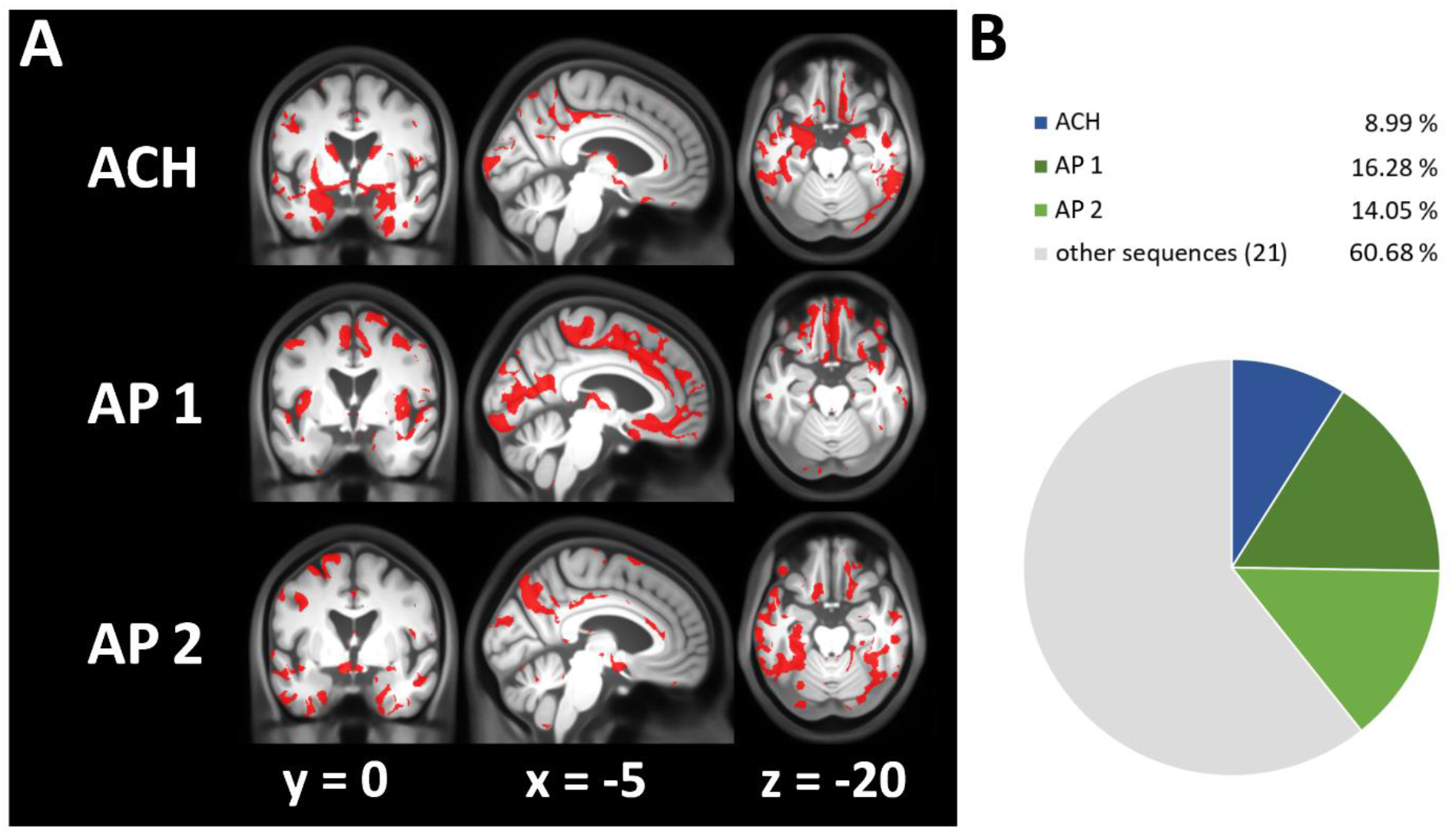
Face validity of ACH using VBM. Voxel-based evidence for monotonic volume decline over 24 sequences gained by permutation of the ACH sequence (ACH, A-T-N-→A+T-N-→A+T+N-→A+T+N+); AP 1: A+T-N-→A+T+N-→A-T-N-→A+T+N+; AP 2: A+T-N-→A-T-N-→A+T+N-→A+T+N+; A: voxels where sequence shows highest evidence; B: percentage of gray matter voxels where sequence has highest evidence.

However, this analysis also suggested that the ACH sequence was not the most evident progression (among 24 tested) in frontal lobe, insular cortex, precentral and postcentral gyri or the cerebellum. Our analysis revealed several brain regions in which alternative sequences over ATN groups were better reflective of monotonic volumetric decline. More specifically, alternative progressions AP 1 (A+T-N-→A+T+N-→A-T-N-→A+T+N+) and AP 2 (A+T-N-→A-T-N-→A+T+N-→A+T+N+) showed highest evidence in 16.28% and 14.05% of the GM respectively (Figure 5). Interestingly, both assume a transient volume increase when transitioning to Amyloid-positivity (i.e. GM volume of A+T-N-> A-T-N-) followed by the lowest GM volume in A+T+N+. AP 1 was the conversion sequence having highest evidence in cortical regions (especially frontal lobe, orbital frontal, premotor regions, insular cortex). AP 2 showed highest evidence in parts of the posterior MTL, the middle and posterior cingulate gyrus, and cortical clusters (the precuneus, temporal, parietooccipital lobe).

Table 2 shows the results for a similar but complementary ROI-level analysis of monotonic GM decline in amygdala, hippocampus, entorhinal cortex and precuneus defined using an MRI atlas. The ACH-based progression was found to be the best fitting sequence to describe monotonic GM volume loss in amygdala and entorhinal cortex. For all ROIs, the ACH progression was found to be among top 5 most likely sequences (out of 24). Surprisingly, AP 1 showed highest evidence for hippocampal ROI volume loss, while the precuneus volume was best described by AP 2. One potential disadvantage of the definition of the ‘N’ category is the dependence on atlas-based ROIs, e.g. for the hippocampus. When using CSF Total Tau for definition of the ‘N’ category, the ACH sequence was also found to optimally describe monotonic GM loss especially in the MTL. On ROI level the ACH sequence was always the most or second most evident pathway (out of 24; Supplemental File 3&4).

**Table 2:** Face validity of ACH in selected ROIs.

| Region | Sequence with highest evidence | T | P | ACH on rank nr. (of 24) | ACH VX% |
| --- | --- | --- | --- | --- | --- |
| AMG | ACH | -13.69 | 2.53e-34 | 1 | 80.99 |
| Hippocampus | AP 1 | -14.88 | 6.76e-39 | 5 | 26.27 |
| Entorhinal Cortex | ACH | -7.30 | 1.13e-12 | 1 | 48.46 |
| Precuneus | AP 2 | -3.90 | 5.76e-05 | 4 | 13.78 |
| Braak I/II | ACH | -16.11 | 1.15e-43 | 1 | 25.13 |
| Braak III/IV | Other1 | -6.97 | 8.59e-12 | 5 | 17.97 |
| Braak V/VI | Other1 | 5.06 | 3.54e-07 | 5 | 7.31 |

ROI based testing for monotonic volume decline over 24 sequences gained by permutation of the ACH sequence. Braak stage volumes are aggregated Freesurfer ROI volumes. ACH: A-T-N-→A+T-N-→A+T+N-→A+T+N+; AP 1: A+T-N-→A+T+N-→A-T-N-→A+T+N+; AP 2: A+T-N-→A-T-N-→A+T+N-→A+T+N+;
*Other1: A+T+N- → A+T-N- → A-T-N- → A+T+N+. ACH VX%: Percentage of voxels with highest evidence* *for ACH sequence inside the ROI mask.*

### Comparing progression sequences towards AD pathology including non-AD continuum groups

All above comparisons were focused on only four ATN groups from the AD continuum (A-T-N-, A+T-N-, A+T+N-, A+T+N+). However, these AD continuum-related groups do not enable direct comparisons of ACH-implied conversion sequences against an alternative timing of events such as Tau positivity preceding Amyloid positivity (e.g. A-T+N+ converting to A+T+N+). We therefore compared the ACH-based sequence to five other biologically possible conversion schemes from A-T-N-towards A+T+N+ (denoted as ANT, TAN, TNA, NAT, NTA). For this comparison the conversion sequences are denoted in the order of each biomarker becoming positive, e.g., TAN stands for: Tau category becomes positive first, Amyloid second, Neurodegeneration last (A-T-N-→ A-T+N-→ A+T+N-→ A+T+N+). Again, Bayesian Information Criterion (BIC) was used to identify conversion sequences with highest evidence for monotonic volume decline both on a voxel as well as ROI-level.

Brain regions with highest evidence for above progressions are characterized in Figure 6. According to our analysis A-first sequences (ACH/ATN, ANT), T-first sequences (TAN, TNA) and N-first sequences (NAT, NTA) showed the highest evidence for monotonic volume decline in 64%, 35% and 0.01% of GM respectively. Local GM regions with highest likelihood for ACH were especially found in the MTL (with an exception of the right anterior hippocampus and parahippocampal gyrus), but also in the basal ganglia (caudate ncl., putamen, thalamus) and precuneus. More supporting clusters for ACH/ATN were observed in all cortical lobes making it clearly the most likely sequence in large parts of GM in this comparison. The ANT progression showed highest evidence in complementary regions of the MTL not covered by ACH/ATN (see above) with additional regions in the basal ganglia (ncl. accumbens), medial frontal lobe, the insular cortex and premotor regions. T-first sequences were most likely only in the cerebellum and some cortical regions including the medial occipital lobe. All of these four sequences showed evidence for different parts of the cingulate gyrus. N-first sequences were only seen in very minor portions of the GM.

**Figure 6:**
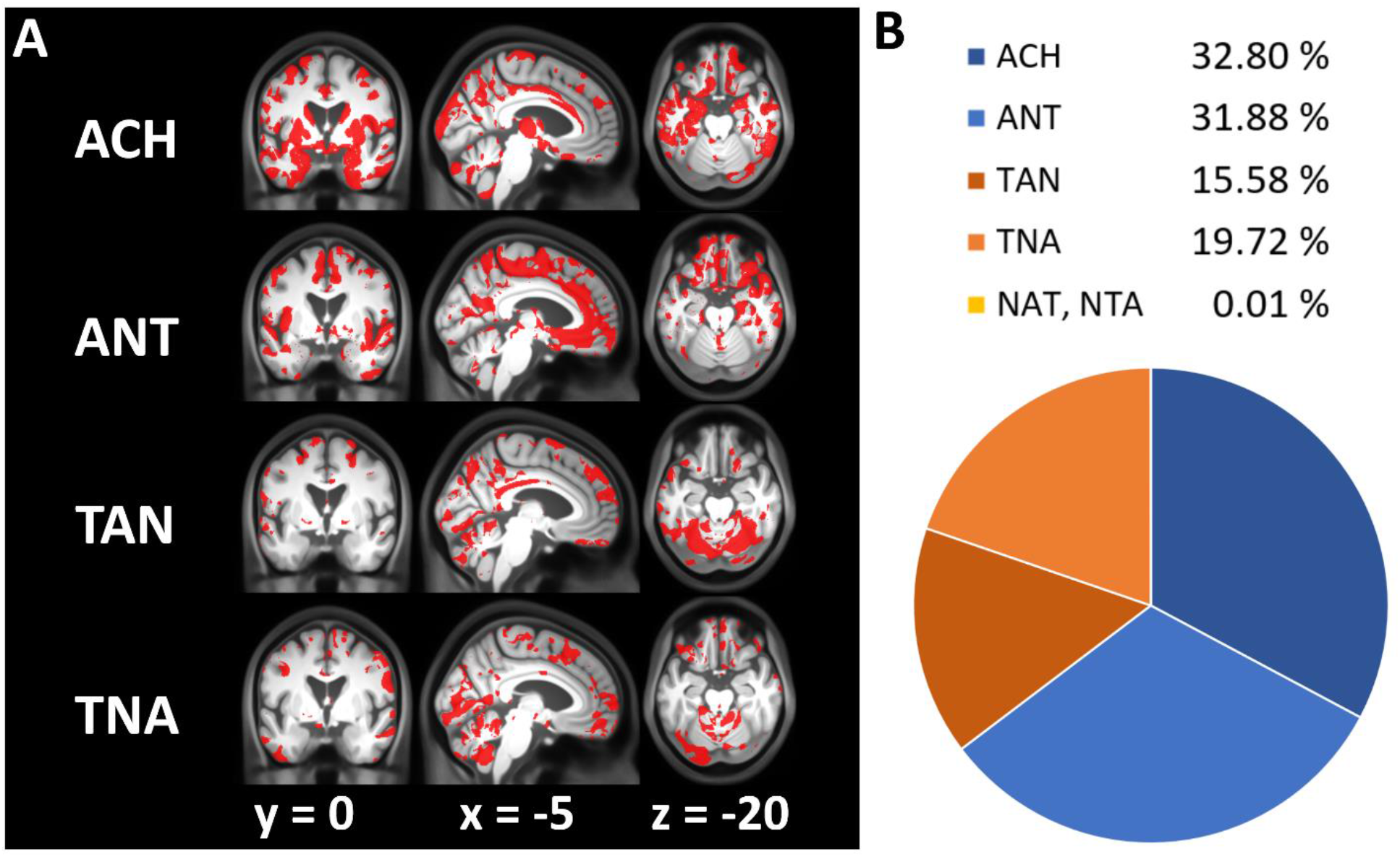
Comparing progression sequences towards AD pathology using VBM. Regions with highest evidence for monotonic volume decline assuming 6 potential disease progressions from A-T-N-towards A+T+N+ (ACH, ANT, TAN, TNA, NAT, NTA). Sequences are denoted in the order of biomarker positivity along the pathway (e.g. ANT = Amyloid-positivity first, Neurodegeneration second, Tau last). A: voxels where sequence shows highest evidence; Notably, regions of highest evidence for each progression are disjunct. B: percentage of gray matter voxels where sequence has highest evidence. N-first sequences (NAT, NTA) are not shown as only few voxels are supported.

In the complementary ROI analysis, conversion sequences with monotonic volume decline were compared for same ROIs as above. The most evident sequences for volume loss per ROI and matching effect size are presented in Table 3. Interestingly, ACH/ATN was the most evident progression for amygdala, entorhinal cortex and precuneus. In hippocampus however, the ANT progression was found to show the highest evidence for the data. Our analysis revealed no indications for the superiority of T-first or N-first over A-first sequences in these ROIs. Notably, above findings were mainly reproduced using CSF Total Tau as neurodegeneration marker. Here, ACH was also the most prominent sequence: On voxel-level, more than 41% of GM showed highest evidence for the ACH sequence (Supplemental File 5). On ROI-level, ACH was the most evident sequence with exception of amygdala and entorhinal cortex, where TAN was more likely which is in contrast to the results of aHV-based Neurodegeneration (Supplemental File 6).

**Table 3:**
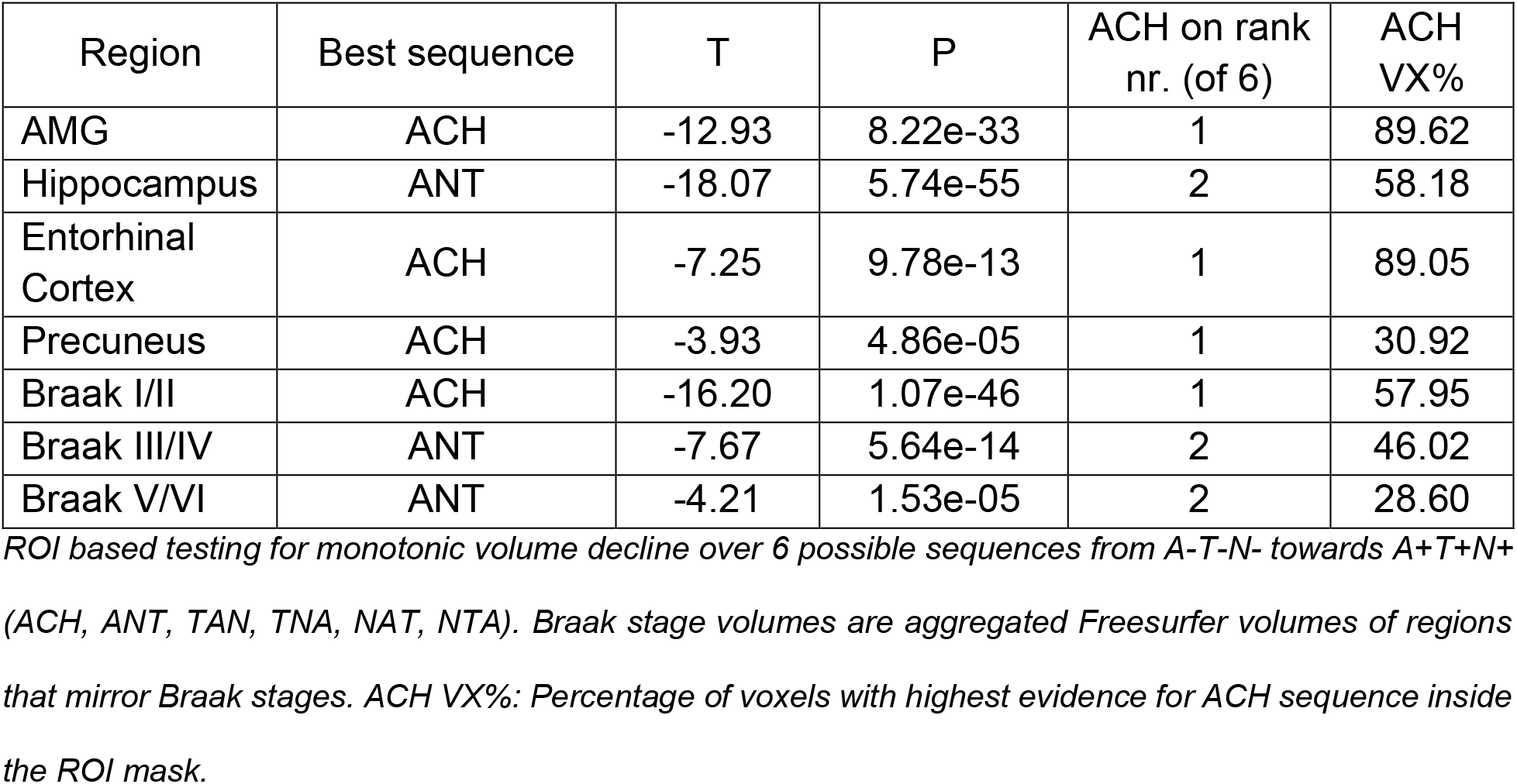
Comparing progression sequences towards AD pathology including non-AD continuum groups in selected ROIs.

| Region | Best sequence | T | P | ACH on rank nr. (of 6) | ACH VX% |
| --- | --- | --- | --- | --- | --- |
| AMG | ACH | -12.93 | 8.22e-33 | 1 | 89.62 |
| Hippocampus | ANT | -18.07 | 5.74e-55 | 2 | 58.18 |
| Entorhinal Cortex | ACH | -7.25 | 9.78e-13 | 1 | 89.05 |
| Precuneus | ACH | -3.93 | 4.86e-05 | 1 | 30.92 |
| Braak I/II | ACH | -16.20 | 1.07e-46 | 1 | 57.95 |
| Braak III/IV | ANT | -7.67 | 5.64e-14 | 2 | 46.02 |
| Braak V/VI | ANT | -4.21 | 1.53e-05 | 2 | 28.60 |

ROI based testing for monotonic volume decline over 6 possible sequences from A-T-N- towards A+T+N+ (ACH, ANT, TAN, TNA, NAT, NTA). Braak stage volumes are aggregated Freesurfer volumes of regions that mirror Braak stages. ACH VX%: Percentage of voxels with highest evidence for ACH sequence inside the ROI mask.

### Concordance of Braak stage trajectory and the ACH trajectory

In addition to above reported ROIs, the two voxel-based analyses of compatibility of the ACH sequence with monotonic GM volume decline were aggregated for larger Braak stage composite regions (Table 2 & 3). Here, stage I/II encompasses the hippocampus and entorhinal cortex, stage III/IV the limbic regions and stage V/VI the remaining cortical regions like precuneus or primary sensory/motor regions. We asked, how much of GM at some stage showed highest evidence for ACH. On voxel-level, the percentage of voxels with highest evidence for the ACH sequence decreased from 25.1% in stage I/II to 7.3% in stage V/VI for the first comparison (face validity of ACH). For the second comparison (including non-AD continuum groups) there was also a noticeable decrease of ACH compatible GM voxels from 58.0% in stage I/II to 28.6% in stage V/VI (see Table 2 & 3).

Thus, on ROI-level, ACH-related ATN sequence across groups was the most evident conversion sequence which is compatible with a monotonic volume decline in Braak stage I/II. For ROIs reflecting stages III/IV and V/VI, ACH was under the most evident 5 (of 24) respectively 2 (of 6) sequences and thus not the most likely explanation for decline anymore. Test statistics and p-values were decreasing with higher Braak stages (stage I/II & III/IV: p < .001; V/VI: p < .05) which supports that ACH compatibility reduces with Braak stage. Using CSF Total Tau instead of hippocampal volume, a similar trend was observed while more voxels supported ACH in stage I/II (stage I/II & III/IV: p < .001; V/VI: p > .05), Supplemental File 3&6).

## Discussion

Since the ATN classification was postulated in 2016^24^, several studies compared ATN groups to a traditional clinical dementia classification^26,27,59,30,29,60,61^. However, a discussion of ATN in context of biological or structural brain changes during AD including local (e.g. voxel-based) brain morphology and the Amyloid Cascade Hypothesis (ACH) can be found to a much lesser extent in previous research^31-33^. This study aimed to focus on these neglected aspects of interest.

Our comparison between ATN status and clinical diagnosis suggested advanced clinical diagnostic status with increasing pathology levels following the ACH group conversion sequence. This is consistent with earlier work^27,59,30^ where a similar pattern has been observed. The A+T+N+ group has previously been reported to show the highest conversion rate to DAT and an increased risk for cognitive decline^26,27,29,60,30,61^. In line with these findings our results suggested substantial memory performance reductions in later ATN stages.

Using MRI and voxel-based morphometry (VBM) we first tested the face validity of a monotonic GM volume decline over the 4 ATN stages (1) A-T-N-(2) A+T-N-(3) A+T+N-(4) A+T+N+ as implied by the ACH and previous work suggesting GM volume loss during ACH progression in clinical DAT. In contrast to previous with strong prior assumptions about ROIs, our emphasize here was on reporting also complementary voxel-based results. More specifically, in line with previous findings brain regions following monotonic volume decline over ATN stages (1)-(4) involved the hippocampus^62,17,46,45,63-68^, amygdala^62,17,46,45,65,67,68^, temporal gyri^17,46,64,66,67^, thalamus^62,17,65,63^, precuneus^62,17,64,66^ and cingulate gyrus^62,17,63,64,66,69^. Consistent with some previous work^69,63,66^ a decline in parts of the cingulate gyrus and insula were observed in our study. Regions that are expected to be affected in very late AD such as frontal^17,66^ and occipital^64,45,63,66^ lobes or less affected such as those around central sulcus^46,64,66^ showed only minor effects in terms of a monotonic decline over ATN stages (1)-(4).

Notably the ACH is still part of an ongoing discussion as anti-amyloid treatments showed only limited success^70,71^. Furthermore, the causal link between Amyloid, Tau and resulting neurodegeneration and dementia is a challenging research topic^19,20^ which might be mediated by alterations in neuropil^71^, synapses^72^ or functional connectivity^73^. Our study revealed strong evidence for an ACH-related monotonic atrophy pattern both on voxel-level but also on ROI-level especially focussed on typical AD-related regions such as the MTL. However, we also observed many gray matter areas where a monotonic volume trajectory along the ACH-implied group progression sequence did show the highest evidence. We identified alternative progressions (denoted AP 1 and AP 2) indicating that there are several cortical regions where the GM volume was found to be higher in the A+T-N-group than in A-T-N-. These were mainly found in cortical regions which are often less strongly affected by AD^66,17^. One might speculate that either Amyloid deposition related tissue expansion^74,75^ and/or sampling effects due to individual differences of brain reserve^76^ might alter brain volume patterns across ATN groups and the progression along the ACH trajectory. A biphasic model of neurodegeneration has been previously suggested by Fortea et al.^74^ hypothesizing that cortical thickening might occur when Aβ becomes abnormal, which presumably reflects inflammation-related swelling, followed by thinning once tau pathology emerges. The authors found Amyloid-related thickness increases in middle temporal, inferior and superior parietal, occipital regions and precuneus which is similar to our findings. Another recent publication also observed Amyloid-related regional volume increase in A+T-N-for the basal forebrain, postcentral gyrus, middle occipital gyrus, and putamen when comparing to A-T-N-^77^. As described above, large parts of the cingulate gyrus did suggest a monotonic volume decline along ACH group progression. However, ACH did not necessarily reveal the highest evidence in the entire cingulate as there were also portions with highest evidence for non-monotonic volume decline (subcallosal to middle cingulate in AP1 & middle to posterior cingulate in AP2). This renders the cingulate a potential candidate region for brain reserve or biphasic model that requires further research.

Under the assumption that patients will convert from negative to positive ATN biomarkers, six patterns of conversions are possible when non-AD pathology ATN groups are additionally included. When testing for monotonic volume decline across these patterns, the highest evidence was found for sequences where Amyloid converts before either Tau or Neurodegeneration (e.g. ATN, ANT), and this was observed in 65% of all gray matter brain areas, especially in AD-related regions. We were able to replicate our finding using ROI-based analyses. In contrast, conversion patterns where Tau converted before Amyloid (TAN, TNA) showed highest evidence for a monotonic volume decline in cortical regions that are atypical for AD pathology. Support for our findings comes from a longitudinal study of 262 non-demented elderly to monitor ATN biomarker progress^32^. It was found that ACH was the most common path of biomarker conversion, but also ANT, TAN and NAT occurred. In contrast, we observed no evidence for NAT in terms of GM volume decreases. It is worth mentioning that per definition, A-T+ or A-N+ groups are not part of the AD-continuum, these groups might initially point to other diseases like primary tauopathies, hippocampal sclerosis/TDP-43 or ischaemic diseases^24^. As remarked by ^1^ and ^32^, the occurrence of conversion sequences other than ACH in real world data might be explained (a) by coincidence of AD- and non-AD pathologic changes (e.g. in A+T-N+) or (b) long-time subthreshold biomarker trends matching the ACH that are not recognized due to a binary classification with disadvantageous thresholds^1,32^. Another study provides more support for our hypothesis of an ACH-related temporal order of biomarker progress by monitoring between-group biomarker changes in an longitudinal approach using the ATN classification^31^.

Furthermore, our morphometric study revealed evidence to support the consistency between the ACH and Braak staging. Brain regions that are expected to be earlier affected by AD-pathology-linked tau deposition (stage I-IV, MTL, limbic system) showed stronger evidence for monotonic GM volume decline over a sequence of conversions than later stages (stage V/VI, cortical regions). As it is already known that brain atrophy often follows Tau and NFT aggregation^65^, both hypotheses were not mutually exclusive. In our analysis, the stronger evidence for volume decline in the amygdala was surprising when comparing to the hippocampus. As ^78^ remarked, Tau pathology in the amygdala is already beginning with Braak stage I/II. However, this effect was not reproducible with Neurodegeneration by CSF Total Tau.

It is known that alternative choices of markers for the N category may have a strong impact on ATN status assignment and longitudinal prediction of cognition^79,80^. The large pool of possible classification methods limits intercomparability between ATN studies dramatically. In our large study, both variants of ATN classification approaches showed converging evidence for the ACH hypothesis. We were not able to determine a superior combination, as both tested N markers have advantages and caveats. The usage of aHV leads to overall stronger effect sizes but one might argue that there is a circularity in defining ATN groups using volumetry and analyzing ATN-related local brain morphometry. Neurodegeneration defined by aHV is a discrete marker of general neuronal loss and the group assignment was carried out using a ROI-rather than voxel-based approach. GM volume (as analyzed by VBM) on the other hand allows a continuous whole brain local analysis on voxel-level and does not align with N for most brain regions.

Although VBM revealed the highest evidence for ACH-related monotonic volume decline in the hippocampus, regions in cortical areas showed compatible monotonic progressions. Taking advantage of the voxel-based approach, differences between sequences could be identified even inside the hippocampus (ACH supports anterior Hippocampus, AP2 posterior hippocampus). It is worth mentioning that the approach is not limited to VBM measurements, in contrast to developmental and plasticity studies stronger expectations for monotonic trajectories do exist for brain volumes in aging and AD. Alternative N categories (such as CSF Total Tau) might also have limitations. The combination of CSF phospho-Tau (T category) and CSF Total Tau (N category) is used in some studies^26,60,61^, while a strong correlation between both markers strongly underrepresents some ATN groups (A?T+N-, A?T-N+). In a recent publication^28^ it was possible to replace CSF Total Tau by CSF phospho-Tau without significant impact on the model. Although there is only a weak correlation between aHV and CSF Total Tau as N markers in our study, a similar pattern of local GM volume decline was revealed. This further suggests that a morphometrical analysis with aHV is applicable.

## Limitations

This study has several methodological limitations. The first challenge was the hippocampal cut-off estimation: The large sample size does not allow to perform atrophy reference methods like autopsy or visual rating of FDG-PET or MRI. Thus, no estimation of sensitivity or specificity was possible, which prohibits a ROC analysis and Youden’s index. Although aHV has a clear unimodal gaussian distribution, it is possible to perform a Gaussian mixture modelling to separate between normal and decreased volumes. A similar approach was performed by^77^. As our data is cross-sectional, no real progression over disease progression and conversions can be modelled and tested. We compensated potential influences of covariates by correcting for demographic marker such as age, sex and education, vascular damage and intracranial volume. This improves comparability (matching) across different ATN groups and increases validity of the underlying cross-sectional progression. Once available, longitudinal DELCODE follow-up data will be used for further validation.

## Conclusion

Early Amyloid status conversion (before Tau and Neurodegeneration) aligns with pattern of brain volume loss observed during AD progression. The ATN classification and the Amyloid cascade hypothesis are compatible with a monotonic progression of MTL atrophy but using the ATN classification system for staging our study revealed indications for non-monotonic progressions in other areas such as several cortical fields.

## Data Availability

The code used during the current study are available from the corresponding author on reasonable request. Data, study protocol and biomaterials can be shared with partners based on individual data- and biomaterial transfer agreements. Requests can be addressed to the DELCODE steering committee

## Glossary

ACH: Amyloid Cascade Hypothesis
AD: Alzheimer’s disease
DAT: Dementia of Alzheimer’s type
aHV: Adjusted hippocampal volume
AN(C)OVA: Analysis of (co-) variance
ATN: Classification based on Amyloid / Tau / Neurodegeneration pathology
BIC: Bayesian Information Criterion
CAT: Computational Anatomy Toolbox
CDR: Clinical Dementia Rating
CERAD: Consortium to establish a registry for Alzheimer’s disease
NC: Non-complaining healthy control
CSF: Cerebrospinal fluid
FDR: False Discovery Rate
FLAIR: Fluid attenuated inversion recovery
GM: Gray matter
MCI: Mild Cognitive Impairment
MMSE: Mini-mental state examination
MPRAGE: Magnetization prepared rapid gradient echo
MRI: Magnetic resonance imaging
MTL: Medial temporal lobe
NFT: Neurofibrillary tangles
ROI: Region of interest
SCD: Subjective Cognitive Decline
SPM: Statistical Parametric Mapping
TICV: Total intracranial volume
VBM: Voxel-based Morphometry
WM: White matter
WMH: White matter hyperintensities

## Declarations

### Ethics approval and consent to participate

DELCODE is retrospectively registered at the German Clinical Trials Register (DRKS00007966), (04/05/2015) and was approved by ethical committees and local review boards. All participants gave written informed consent to participate.

### Consent for publication

Not applicable

### Author’s contributions

The study was designed by E.D., M.W., A.S. and F.J. Data analysis was performed by N.H. N.H., G.Z., and E.D. wrote the manuscript. All authors reviewed the paper.

## Acknowledgements

Not applicable

## Supplement

**Supplemental File 1:**
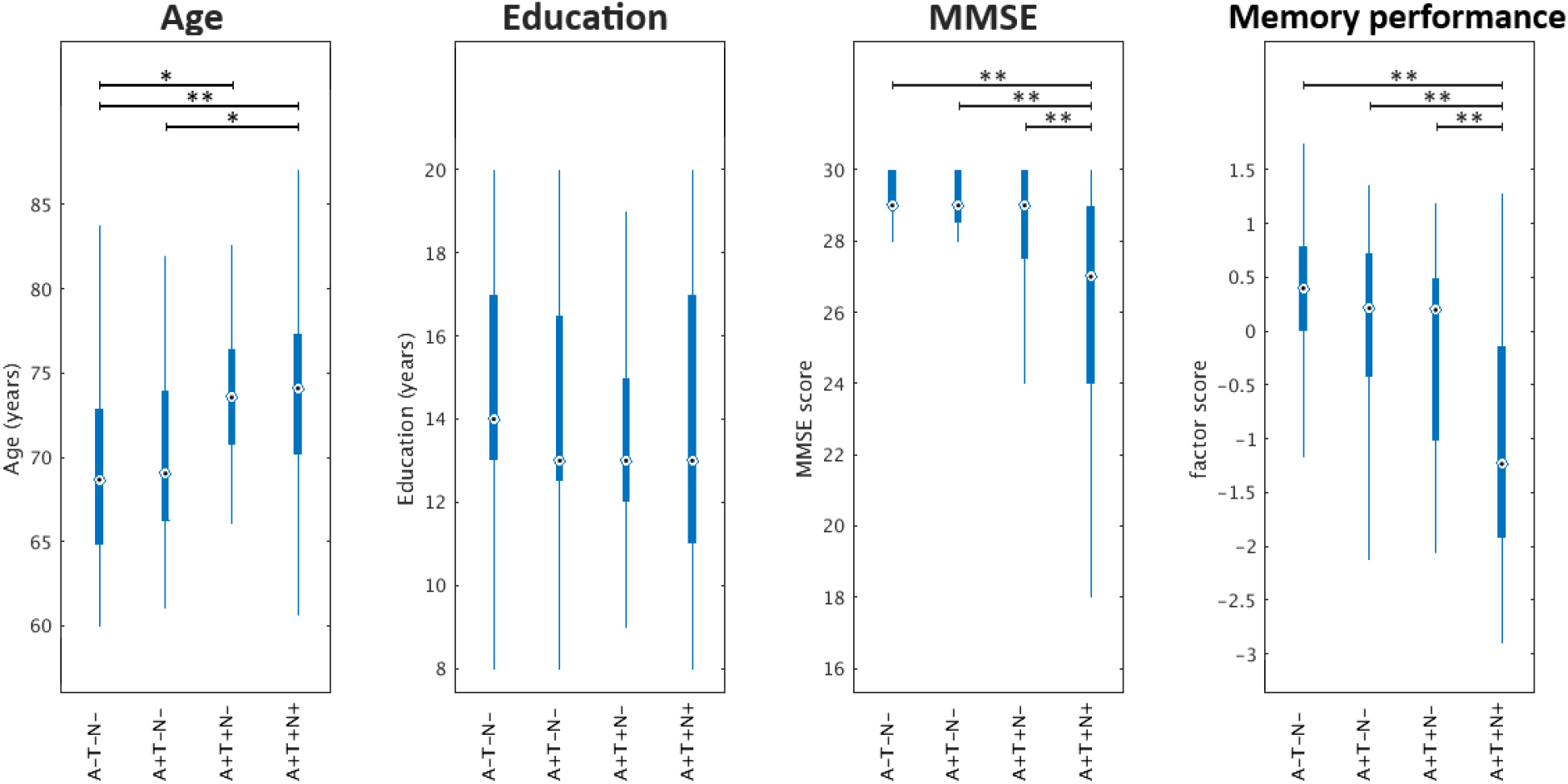
Comparison between selected ATN groups using CSF-total-Tau. Boxplots of age, sex, cognition for selected ATN groups. *: p < .05 after Bonferroni correction, **: p < .001 after Bonferroni correction. Neurodegeneration (N) by CSF Total Tau.

**Supplemental File 2:**
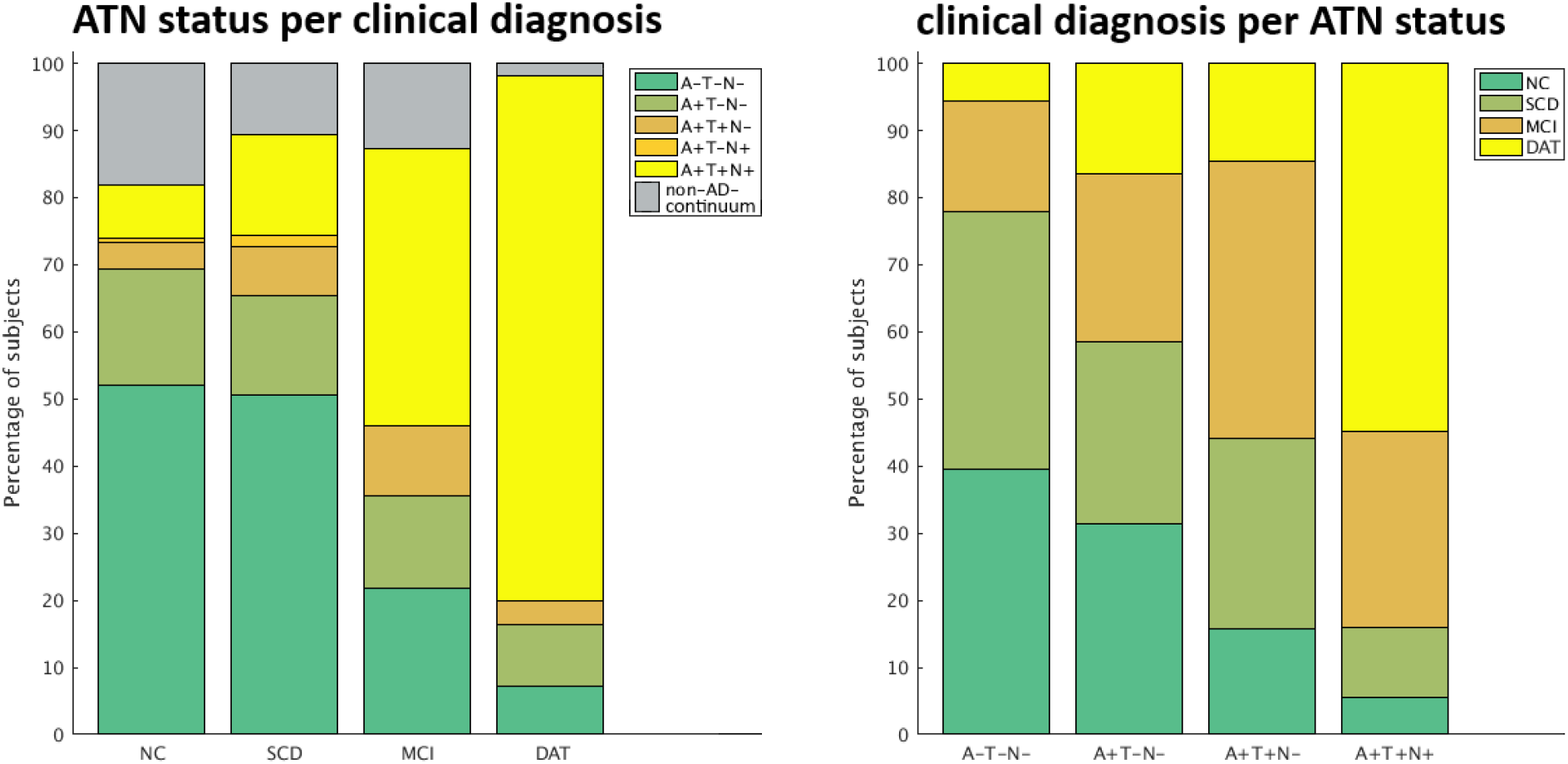
Distribution of ATN status and clinical diagnosis using CSF-total-Tau. Left: percentual distribution of selected ATN groups per clinical diagnosis; right: percentual distribution of clinical diagnosis per ATN groups. Neurodegeneration (N) by CSF Total Tau.

**Supplemental File 3:**
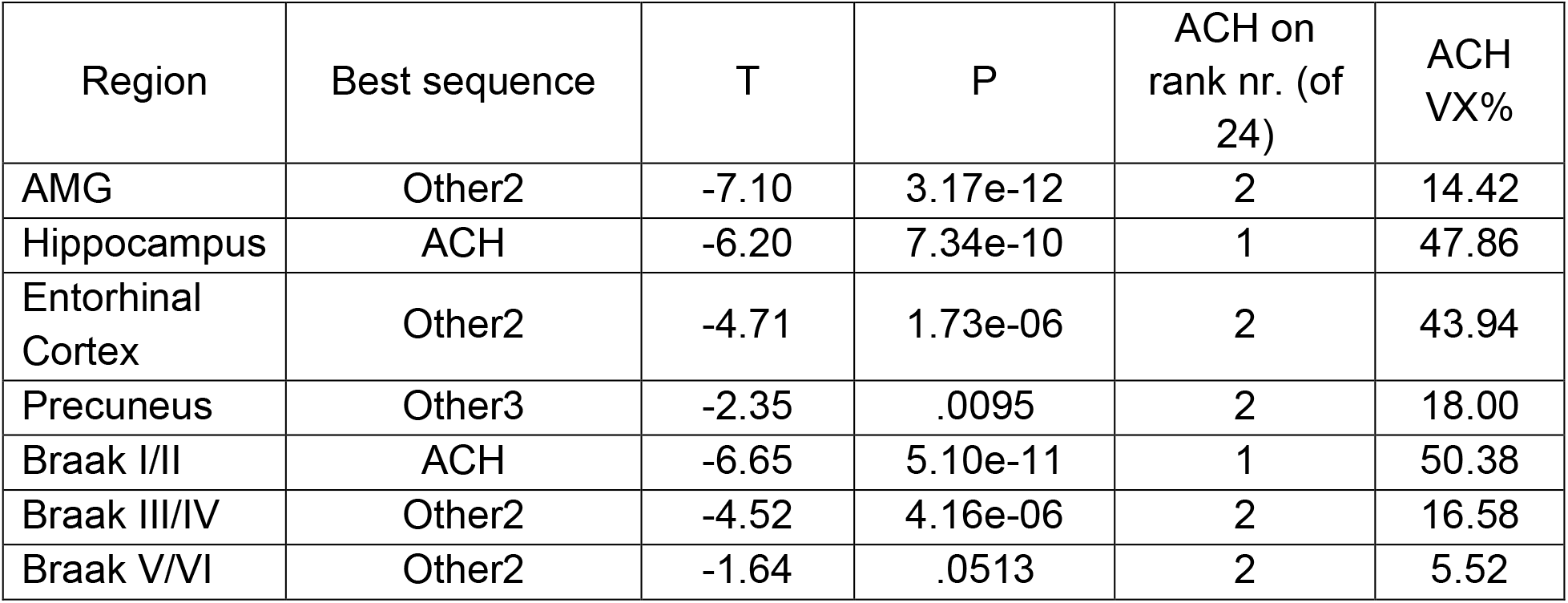
Face validity of ACH using selected ROIs and CSF-total-Tau. ROI based testing for monotonic volume decline over 24 sequences gained by permutation of the ACH sequence. Braak stage volumes are aggregated Freesurfer ROI volumes. Other2: A-T-N-→ A+T+N-→ A+T-N-→ A+T+N+; Other3: A+T-N-→ A-T-N-→ A+T+N-→ A+T+N+. ACH VX%: Percentage of voxels with highest evidence for ACH sequence inside the ROI mask. Neurodegeneration (N) by CSF Total Tau.

**Supplemental File 4:**
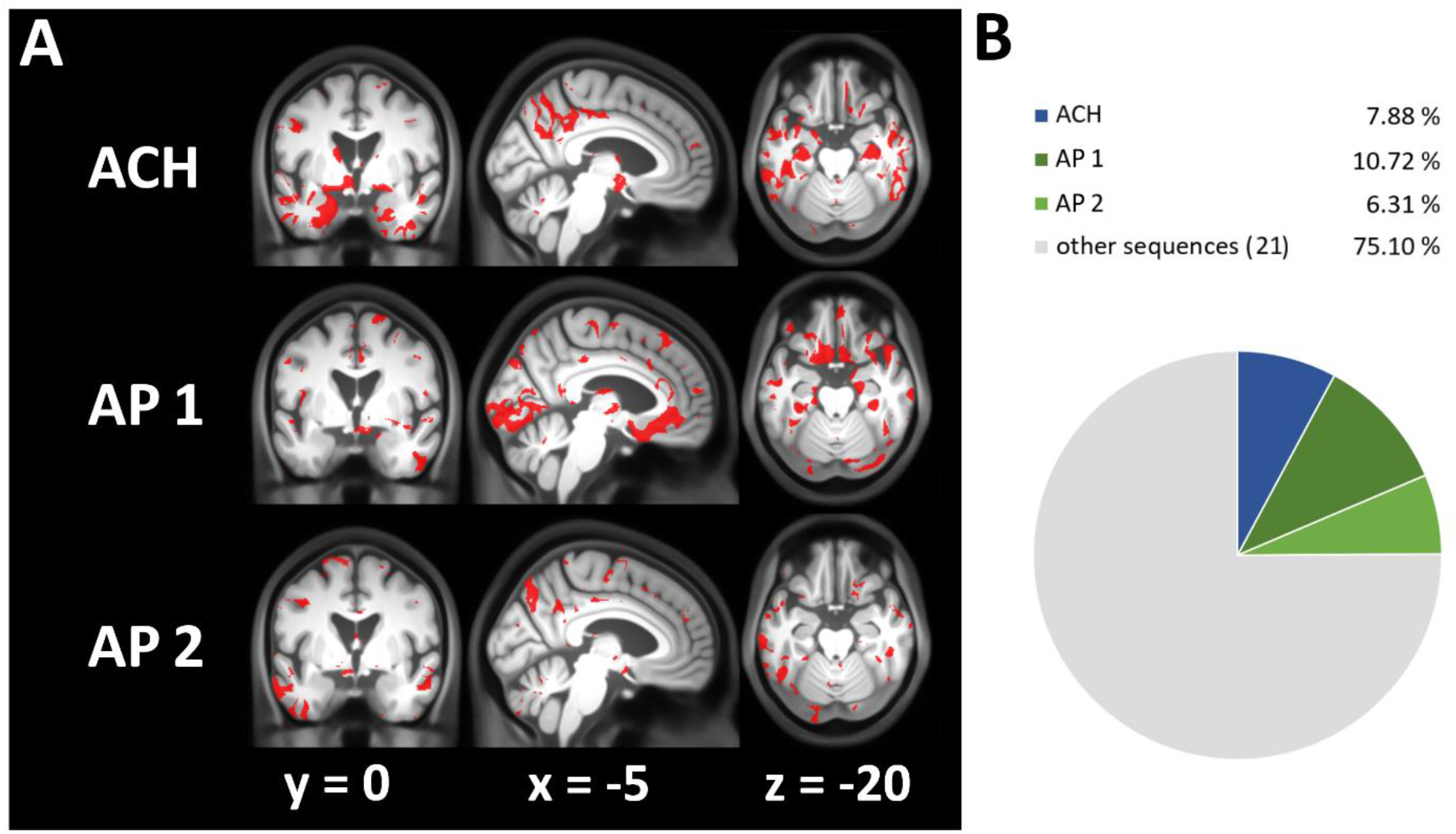
Face validity of ACH using VBM and CSF-total-Tau. Voxel-based evidence for monotonic volume decline over 24 sequences gained by permutation of the ACH sequence (ACH, A-T-N-→A+T-N-→A+T+N-→A+T+N+); AP 1: A+T-N-→A+T+N-→A-T-N-→A+T+N+; AP 2: A+T-N-→A-T-N-→A+T+N-→A+T+N+; A: voxels where sequence shows highest evidence; B: percentage of gray matter voxels where sequence has highest evidence. Neurodegeneration (N) by CSF Total Tau.

**Supplemental File 5:**
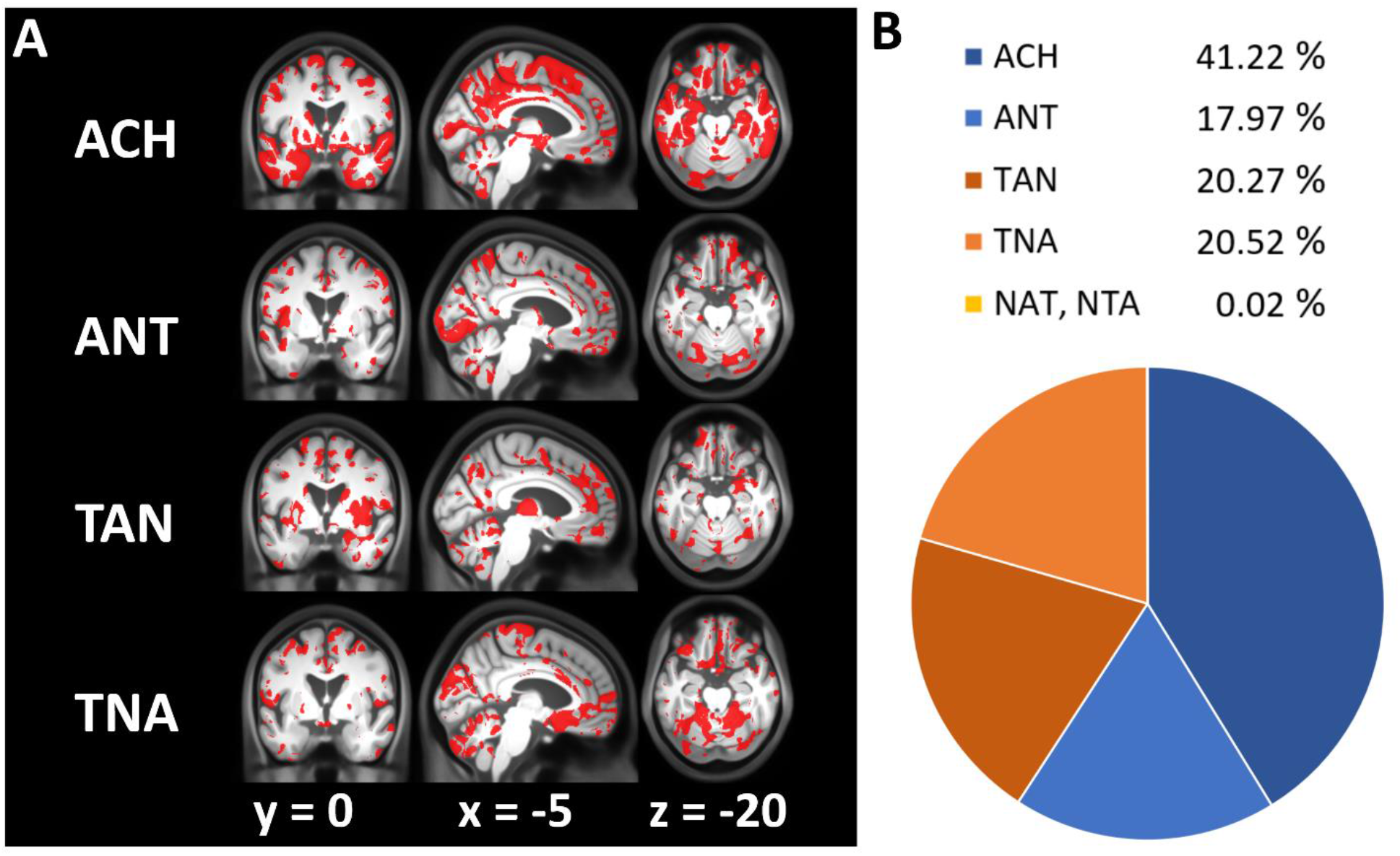
Comparing progression sequences towards AD pathology using VBM and CSF-total-Tau. Voxel-based evidence for monotonic volume decline over 6 possible sequences from A-T-N-towards A+T+N+ (ACH, ANT, TAN, TNA, NAT, NTA). Sequences are denoted in the order of biomarker positivity along the pathway (e.g. ANT = Amyloid-positivity first, Neurodegeneration second, Tau last). A: voxels where sequence shows highest evidence; B: percentage of gray matter voxels where sequence has highest evidence. N-first sequences (NAT, NTA) are not shown as only few voxels are supported. Neurodegeneration (N) by CSF Total Tau.

**Supplemental File 6:**
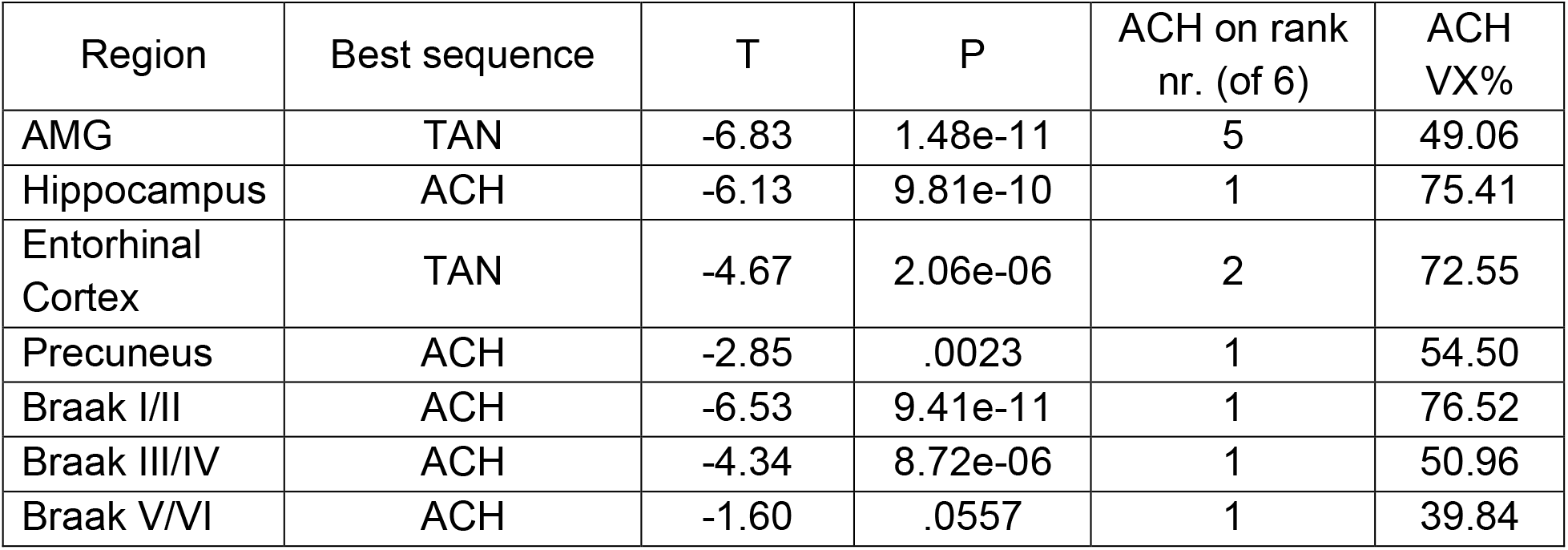
Comparing progression sequences towards AD pathology using selected ROIs and CSF-total-Tau. ROI based testing for monotonic volume decline over 6 possible sequences from A-T-N-towards A+T+N+ (ACH, ANT, TAN, TNA, NAT, NTA) including also non-AD continuum groups. Braak stage volumes are aggregated Freesurfer ROI volumes. ACH VX%: Percentage of voxels with highest evidence for ACH sequence inside the ROI mask. Neurodegeneration (N) by CSF Total Tau.

